# Enhancing Precision Drug Therapy and build pharmacokinetic model in Pregnant Women: PBPK Modeling of Antiviral drugs

**DOI:** 10.1101/2024.07.22.24310817

**Authors:** Mohamed Abdulsamed, Ashraf. A. Naass, Mohamed. S. AEswani, Sedigh Bashir

**Affiliations:** Libyan Biotechnology Research Center

**Keywords:** Pharmacokinetic, Modeling, Pregnancy, Antiviral drugs, PBPK. Maternal-fetal

## Abstract

PBPK/PD modeling is essential in modern drug development. Traditional drug development methods frequently rely on trial and error, which can be time-consuming, costly, and could be risky. Predicting pharmacokinetics (PK) of drugs in pregnant women, encompassing the intricate aspect of placental drug transfer, remains a complex task. This study was to compare of simulated or predicted and observed (previously published approaches) pharmacokinetic parameters among the four antiviral drugs in pregnant and non-pregnant women. In addition, this investigation endeavors to construct and assess physiologically-based pharmacokinetic (PBPK) models specific to maternal-fetal interactions for four antiviral drugs, Acyclovir, Emtricitabine, Dolutegravir (DTG) and Raltegravir (RAL). PBPK models were built with the Open Systems Pharmacology software suite (PK-Sim/MoBi). Different approaches to inform placental drug transfer were applied and compared. Model performance was evaluated using in vivo all 4 a forementioned antiviral maternal plasma concentrations during the 2nd and 3rd trimesters and umbilical vein concentrations at delivery. All clinical in vivo data were obtained from the International Maternal paediatric and Adolescent AIDS Clinical Trials (IMPAACT) Network P1026s study. The PBPK models successfully predicted plasma concentration-time profiles of four antiviral drugs in the 2nd and 3rd trimesters and most predicted PK parameters fell within a 1.33-fold error range. Predicted umbilical vein concentrations of DTG among others were in reasonable agreement with in vivo data but were sensitive to changes in the placental partition coefficient and transplacental clearance. Maternal-fetal PBPK modeling reliably predicted maternal PK of previously mentioned antiviral during pregnancy. For the fetal PK, data on the unbound fraction of highly protein-bound DTG has proven to be important to adequately capture changes in total clearance in silico. More research efforts, along with clinical data, are needed to verify the predictions of fetal PK of antiviral. In conclusion, the findings suggest the feasibility of employing physiologically-based pharmacokinetic (PBPK) models to assess the disposition of antiviral drugs in pregnant women and their fetuses.

## Introduction

Medication utilization during pregnancy is widespread and increasing. In a prospective, longitudinal cohort study focusing on prescription drugs and other medication consumption during pregnancy, findings indicate that 97.1% of women included in the study engaged in the use of at least one medication throughout their pregnancy, and 30.5% women took 5 or more medications.^1^ Although physiological changes during pregnancy can notably influence drug disposition, the inclusion of pregnant women in clinical studies remains limited. Consequently, clinicians frequently encounter the challenge of prescribing medications during pregnancy without access to specific information regarding pharmacokinetics (PK) and safety in this population. Particularly, antiviral medications are commonly administered during pregnancy for both maternal treatment and prophylaxis to mitigate the risk of perinatal viral transmission. Acyclovir, an antiviral drug effective against the herpes simplex virus, stands as a noteworthy example. Herpes simplex virus constitutes one of the most prevalent sexually transmitted infections, bearing the potential for neonatal morbidity or mortality if infection during the neonatal period is not averted or promptly addressed^2^. Emtricitabine, Dolutegravir, and Raltegravir represent antiretroviral drugs with efficacy against human immunodeficiency virus (HIV) infection. As of 2016, a substantial cohort of 19.5 million individuals living with HIV is actively receiving antiviral treatment^3^. With the expanding depth of knowledge concerning anatomical and physiological alterations during pregnancy, the application of physiologically based pharmacokinetic (PBPK) models to extant pharmacokinetic (PK) data for pregnant women has become feasible, enhancing confidence in these models. Leveraging their mechanistic foundation, PBPK models offer valuable insights into the physiological mechanisms underpinning PK changes. Anticipating PK alterations in specific populations, such as pregnant women, prior to the commencement of clinical trials, can streamline the design and execution of such studies. However, a prerequisite for such applications is a robust confidence level in the established PBPK model. While several PBPK models for pregnancy have been established.^4^ a notable gap persists, as many predominantly focus on maternal PK changes, with limited consideration for drug exposure in the fetal compartment^5^.According to the collected data This study presents the development of maternal-fetal PBPK models for group of antiviral drugs which are acyclovir, Emtricitabine, Dolutegravir and Raltegravir. The objectives of this study were to compare of simulated or predicted and observed (previously published approaches) pharmacokinetic parameters among the four antiviral drugs in pregnant and non-pregnant women and to evaluate the model predictions of the PK in the mother at different stages of pregnancy.

### Methodology

The study was based on data collection through PubMed, Scopus and Google scholar using the medical terms “Physiologically based pharmacokinetic pregnancy modeling” and Antiretroviral drugs. These obtained data and literature were focused on using open systems pharmacology software package version 8.0 (https://www.open-systems-pharmacology.org/). (PK-Sim) and MoBi. In addition, R Foundation for stastical computing version 3.4.1 software. On other hand, all clinical in vivo data were obtained from the international maternal pediatric and adolescent AIDS clinical trial (IMPAACT) Network P 1026s study. All sources code and the model developed available on GitHub accessible via(https://www.open-systems-pharmacology.org/).

### General Workflow

The procedural framework governing the advancement of the pregnancy Physiologically-Based Pharmacokinetic (PBPK) model has been exhaustively explicated in a previous scholarly account^6^. The schematic depiction of this methodology is presented in Figure 1. Briefly, a PBPK model was initially developed for a virtual non-pregnant population and evaluated by comparing simulation results with the observed in vivo PK data in non-pregnant subjects reported in the comparison studies. Thereafter, the non-pregnant PBPK model was translated to pregnancy by substituting the standard model structure with the pregnancy structure and parametrizing the model for the respective gestational age as described before. ^6^PK predictions in pregnant women were evaluated by comparison with in vivo PK data obtained from clinical trials of IMPAACT P1026s.

**Figure 1.**
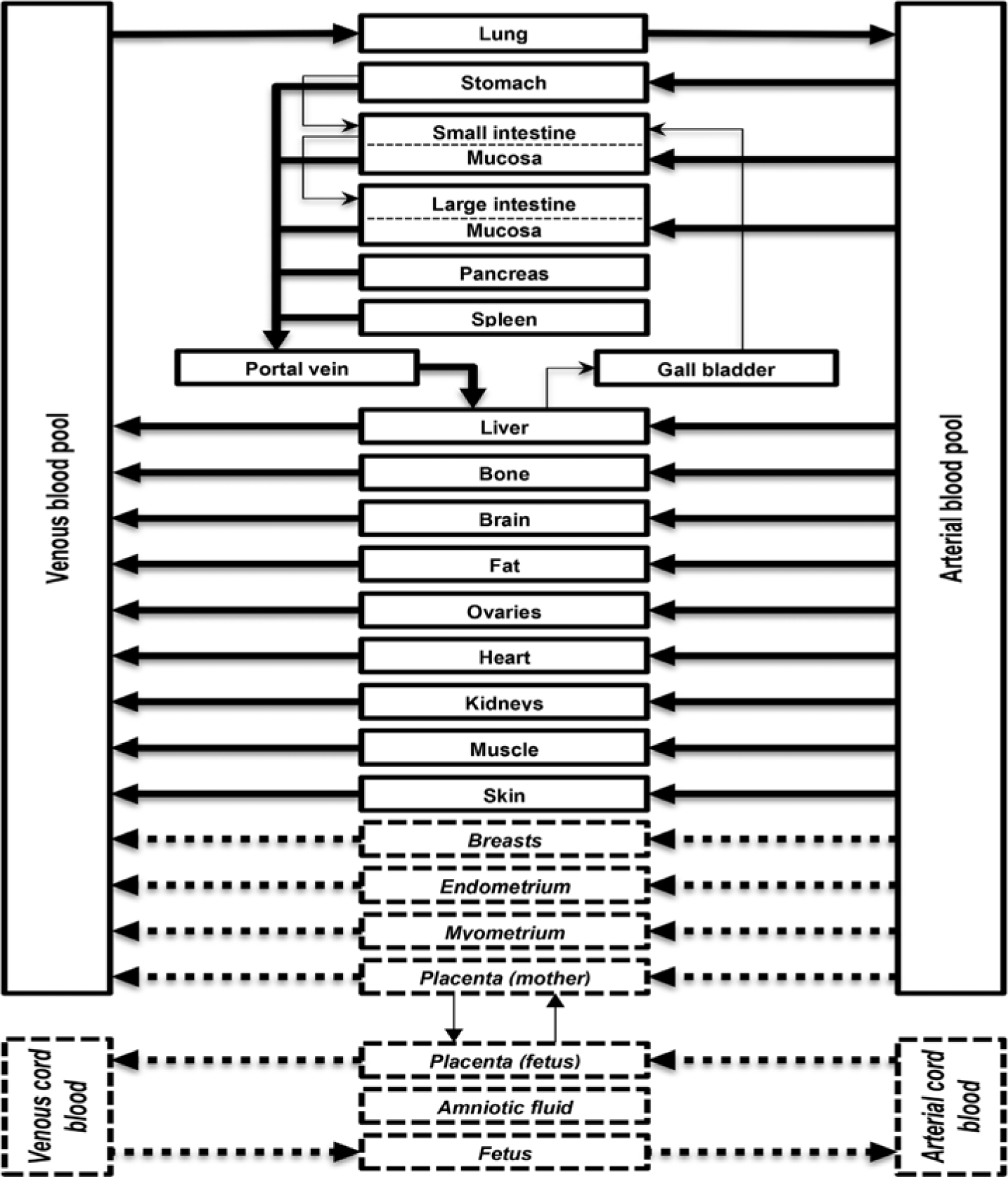
Pregnancy PBPK model structure. Thick arrows represent drug transport via blood flow, and thin arrows via other pathways (eg, via passage in the gastrointestinal tract, biliary excretion through the gallbladder, and diffusive transfer in the placenta). Compartments that are exclusively part of the pregnancy PBPK model structure are shown in italics with dashed borders and dashed arrows for drug transfer via the blood flow.^7^

### Development of PBPK Models

**Acyclovir** demonstrates renal excretion efficiency after intravenous administration, with a range of 61% to 91% of the radioactively labeled dose being excreted unchanged in urine (corrected for the amount of radioactivity lost), and 8.5% to 14.1% is metabolized to CMMG (9-carboxy methoxymethylguanine).^8^ OAT (organic anion transporter) 2 has been suggested to be the main transporter involved in renal secretion.^9^ CMMG is formed in a 2-step reaction involving a reversible oxidation catalyzed by alcohol dehydrogenase^10^ and subsequent irreversible transformation to CMMG via aldehyde dehydrogenase 2.^11^. Pharmacokinetic profiles in nonpregnant subjects were obtained from a study by Laskin et al,^12^who investigated acyclovir disposition after intravenous administration of different doses, and additionally from another study^13^ that investigated acyclovir PK after oral administration of 400 mg acyclovir as either a suspension or a tablet. Two studies, encompassing pregnant women nearing full term, have presented maternal plasma concentration-time data subsequent to the administration of 400 mg acyclovir, both in single and multiple dosage scenarios. ^14,15^ and were used for evaluating the predicted maternal plasma concentrations. Another study reported paired concentration measurements in the maternal plasma and in the umbilical vein obtained at delivery,^16^ which were used for evaluating the predicted concentrations in the umbilical vein blood compartment of the model.

**Emtricitabine** is a nucleoside reverse transcriptase inhibitor with a daily oral dose of 200 mg in both pregnant and non-pregnant adult populations.^17^ Emtricitabine undergoes predominant elimination in an unchanged form through renal excretion, involving a combination of glomerular filtration and tubular secretion, accounting for 71% of the radioactive dose (adjusted for lost radioactivity).^18^ PK simulations in the non-pregnant population were evaluated by comparison with in vivo data obtained from 5 different studies that investigated the PK of Emtricitabin**e** in non-pregnant subjects after single and multiple oral administration of 200 mg.^19-20^ In pregnant women the PK was predicted in different populations, namely in 3 different gestational age groups of non-laboring pregnant women (23-30, 31-35, and 36-42 gestational weeks) and in women in labor between 34 and 39 weeks of gestation. Drug concentrations in the umbilical vein were predicted in the latter group. PK predictions in pregnant populations were evaluated through comparison with some hitherto unpublished and published in vivo data.^21^ The clinical in vivo data were from the IMPAACT (International Maternal Pediatric and Adolescent AIDS Clinical Trials) Network study P1026s.^21^ In this clinical study steady-state PK samples were collected at 0, 1, 2, 4, 6, 8, 12, and 24 hours after dosing. The protocol for this study was approved by the responsible institutional review boards^22^.

**Dolutegravir** is prescribed at a daily dosage of 50 mg in the form of an orally administered tablet, administered once daily to both pregnant and non-pregnant adult patients who are treatment-naïve or treatment-experienced, and lack resistance to integrase strand transfer inhibitors (INSTIs). The predominant elimination pathway for Dolutegravir involves metabolic processes facilitated by various enzymes, namely UGT1A1, UGT1A3, UGT1A9, and CYP3A4, accounting for approximately 51%, 2.8%, 5.5%, and 21% of the administered dose, respectively.^23^ In the developed model, the contribution of UGT1A3 and 1A9 to total glucuronidation was combined into the biotransformation pathway mediated by UGT1A1. Finally, to obtain dose fractions summing up to 1.0, the dose fraction metabolized via UGT1A1 was increased to 0.79 based on the assumption that the reported value (0.51) may be underestimated due to hydrolyzation and back conversion of the glucuronide to DTG in the feces, as discussed elsewhere^23^. In the pregnancy PBPK model, physiologic parameters were adjusted to the respective stage of pregnancy^24^. Additionally, the reference concentrations of UGT1A1 and CYP3A4 (quantifying the concentrations of these enzymes in the model) were increased to reflect induction of these enzymes. Specifically, CYP3A4 reference concentration was increased by a factor of 1.60 in the 2nd and 3rd trimesters and UGT1A1 reference concentration by a factor of 1.75 in the 2nd trimester and 1.92 in the 3rd trimester ^25,26^. The fraction unbound of DTG, averaging 0.0070 in non-pregnant subjects,^27^ was also adjusted based on the albumin concentration measured in the herein investigated study subjects. Specifically, the mean albumin concentration measured in the 2nd trimester, 3rd trimester and 6 – 12 weeks postpartum was 34.4 g/L, 32.8 g/L and 41.4 g/L, respectively. Using a previously presented scaling approach^24^, these measurements resulted in a fraction unbound of 0.0084 and 0.0088 in the 2nd and 3rd trimester, respectively. PK simulations in the non-pregnant population were evaluated by comparison with in vivo data obtained from eight clinical studies reported in the literature that investigated the PK of DTG in a total of 22 different groups of non-pregnant subjects after single and multiple oral administrations of 2 to 100 mg as granule suspension or 50 mg as tablet in fasted or fed state ^28-37^. In pregnant women, the PK was predicted in 2 different gestational age groups of non-laboring pregnant women in the 2nd trimester (median gestational age [range]: 23.5 weeks) ^31-36^ and 3rd trimester (median gestational age [range]: 33 ^40-47^weeks), and in women in labor (median gestational age [range]: 38 ^45-52^weeks). Drug concentrations in the blood plasma of the umbilical vein were predicted in the laboring pregnant women group.

**Raltegravir** is prescribed in either 400 mg twice daily or 1200 mg once daily oral tablet regimens for adult patients, encompassing both pregnant and non-pregnant individuals who are treatment-naive or treatment-experienced. The predominant elimination pathway for raltegravir involves metabolism catalyzed by UGT1A1 and UGT1A9, constituting approximately 70% and 11% of the administered dose, respectively.^37^. Additionally, approximately 9% is eliminated unchanged through the kidneys ^37^. The non-pregnant PBPK model for RAL was obtained from the OSP GitHub repository (https://github.com/Open-SystemsPharmacology/Raltegravir-Model/releases) where an extensive description and evaluation of the model can be found. In the pregnancy PBPK model, physiologic parameters and the reference concentrations of UGT1A1 were adjusted to the respective stage of pregnancy as described above. Since no information on the effect of pregnancy on UGT1A9 could be found, this enzyme was not induced in the presented model. Similar to DTG, the fraction unbound of RAL, averaging 0.17 in non-pregnant adultsm,^38^ was adjusted based on the mean albumin concentration measured in the 2nd trimester, 3rd trimester and 6 – 12 weeks postpartum (34.1 g/L, 32.4 g/L and 41.4 g/L, respectively) resulting in a fraction unbound of 0.198 and 0.206 in the 2nd and 3 rd trimester, respectively. Additional information on model development and translation to pregnancy can be found in the Supplemental Material. In pregnant women, the PK were predicted in 2 different gestational age groups of non-laboring pregnant women in the 2nd trimester (median gestational age [range]: 23.5 31-36 weeks) and the 3rd trimester (median gestational age [range]: 34 41-49 weeks), and in women in labor (median gestational age [range]: 38 47-51 weeks). Drug concentrations in the blood plasma of the umbilical vein were predicted in the laboring pregnant women group.

### Evaluation of PBPK Models

The PBPK models were evaluated through visual comparison of observed in vivo plasma concentration-time profiles with the concentrations simulated in non-pregnant women or predicted in pregnant women. Other visual predictive checks included goodness-of -fit (GOF) plots, in which individual in vivo concentration values, if available, were combined at each time point to geometric mean values. Additionally, simulated or predicted PK parameters were compared with observed PK parameters obtained from the mean in vivo plasma concentration time profiles. Ratios of simulated or predicted to observed PK parameters were also estimated.

## Results

### Non-pregnant and Pregnant PBPK models

#### Acyclovir and Emtricitabine

The outcomes detailed herein exclusively apply to female who are not currently pregnant. The Figure 2 illustrates the Simulated of plasma concentration-time profiles of acyclovir subsequent to intravenous administration. In this Figure, the paragraphs presented below depict the Goodness-of-fit plot for plasma concentrations of acyclovir (upper panels) and Emtricitabine (lower panels) in non-pregnant subjects (left panels) and pregnant women (right panels). The continuous line denotes the line of identity, while the dashed lines delineate the 2-fold error range. In the upper left panel, acyclovir plasma concentrations are presented for non-pregnant subjects, with black circles representing concentrations for the suspension and gray circles signifying concentrations in steady state for the suspension. Meanwhile, in the upper right panel, acyclovir plasma concentrations in pregnant women are illustrated, with black circles indicating concentrations after a single dose and gray circles representing concentrations in steady state. Lower left panel: Emtricitabine plasma concentrations in non-pregnant subjects; black circles indicate the concentrations after a single dose, gray circles indicate concentrations at steady state, gray squares indicate concentrations after a single dose, black squares indicate concentrations after a single dose, and black triangles indicate concentrations at steady state. Lower right panel: Plasma concentrations of Emtricitabine in pregnant women are delineated as follows: black circles denote concentrations in women at gestational age 23-30 weeks, gray circles represent concentrations in women at gestational age 31-35 weeks, and black squares indicate concentrations in women at gestational age 36-42 weeks. **^13-21^**

**Figure 2:**
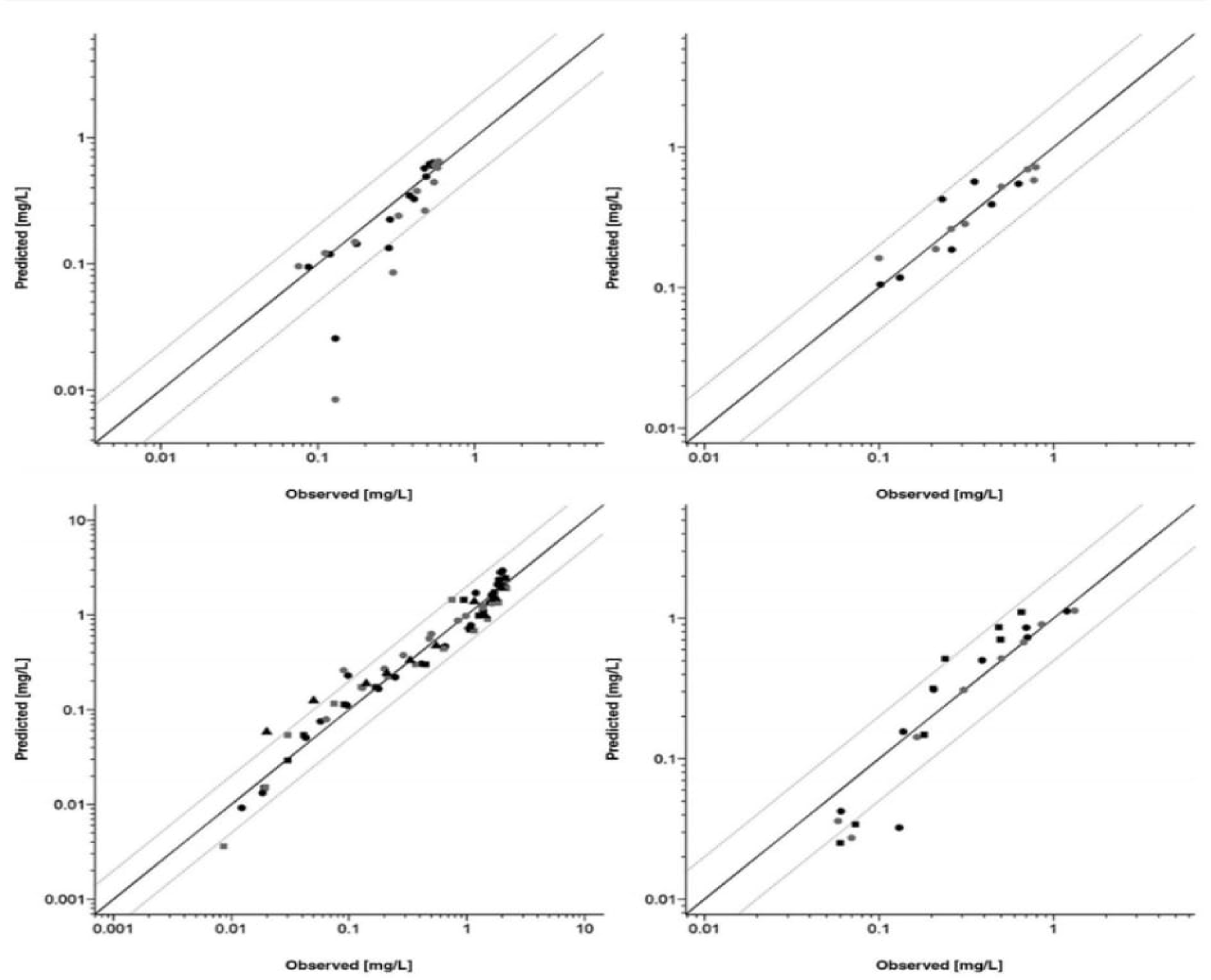
Goodness-of-fit plot for plasma concentrations of acyclovir (upper panels) and Emtricitabine (lower panels) in non-pregnant subjects (left panels) and pregnant women (right panels).

### Evaluation of Predicted Drug Pharmacokinetics for Pregnant Women

The results described in the following refer exclusively to pregnant women. Figure 3 (panels A and B) and Figure 4 (panels A, B, and C) show the anticipated maternal plasma concentration-time profiles of acyclovir and emtricitabine were consistent with the observed in vivo data, demonstrating good agreement between predictions and actual outcomes. Figure 2 presents the predicted mean concentration values in a GOF plot and indicates that all maternal concentrations of acyclovir were predicted within a 2-fold error range, whereas for emtricitabine 79% of the concentration values were predicted within that range. PK parameters calculated from the predicted emtricitabine and acyclovir plasma concentration-time profiles are compared with the observed in vivo PK parameters in Table 2. For acyclovir, the ratios of predicted to observed PK parameters were all within a 25% error range, whereas for emtricitabine most of these ratios were within that range.

**Figure 3.**
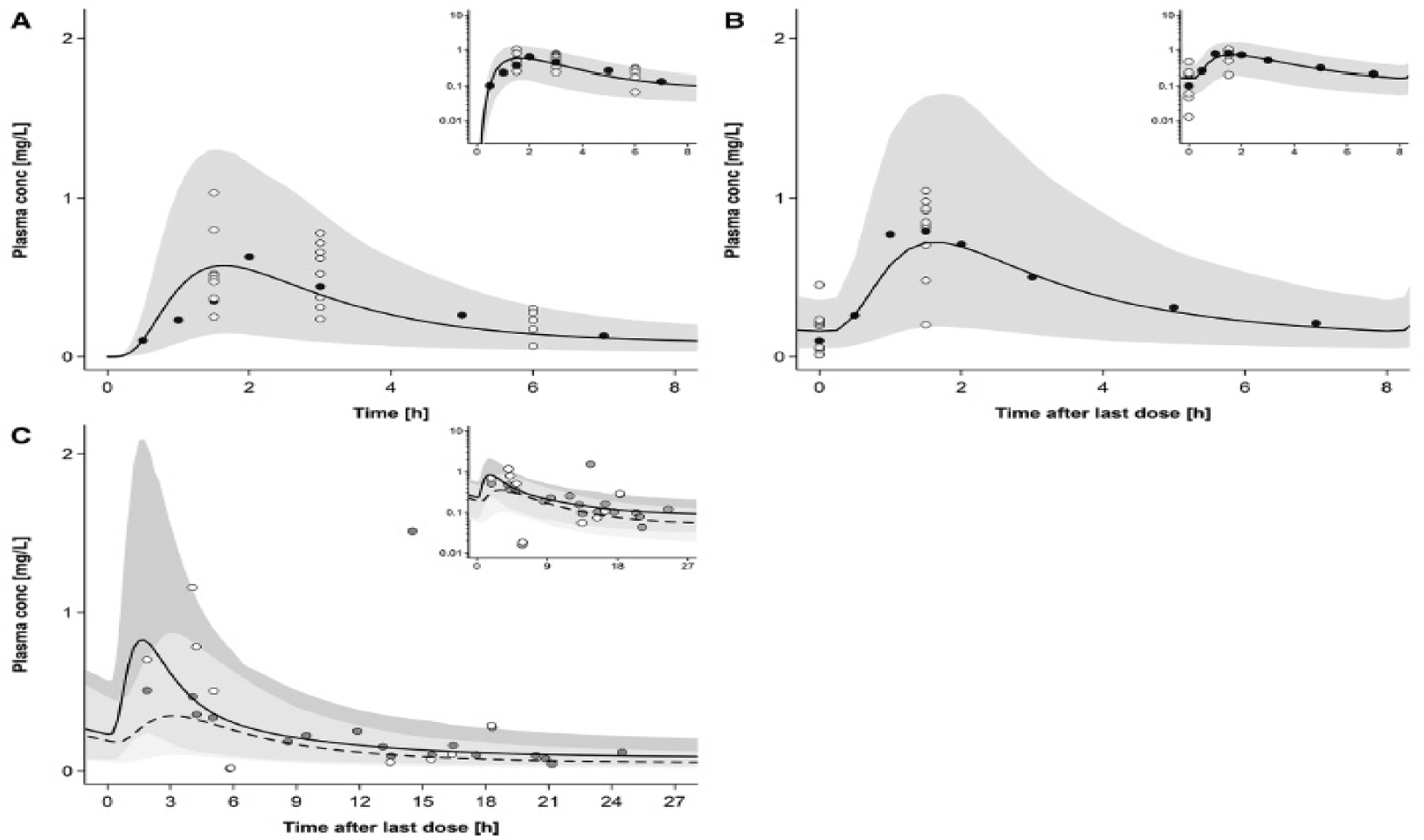
Plasma concentration-time profiles of acyclovir following oral administration of 400 mg in pregnant women. Semi-log scale figures are given as inset figures in the top right corners. Observed in vivo data were taken from published studies. **A graph shows**, Single dose in pregnant women with an average gestational age of 36 weeks. **B graph shows,** Multiple doses in steady state in pregnant women with an average gestational age of 38 weeks. **C graph shows,** Multiple doses in steady state in pregnant women with an average gestational age of 40 weeks.^14,15,16^

**Figure 4.**
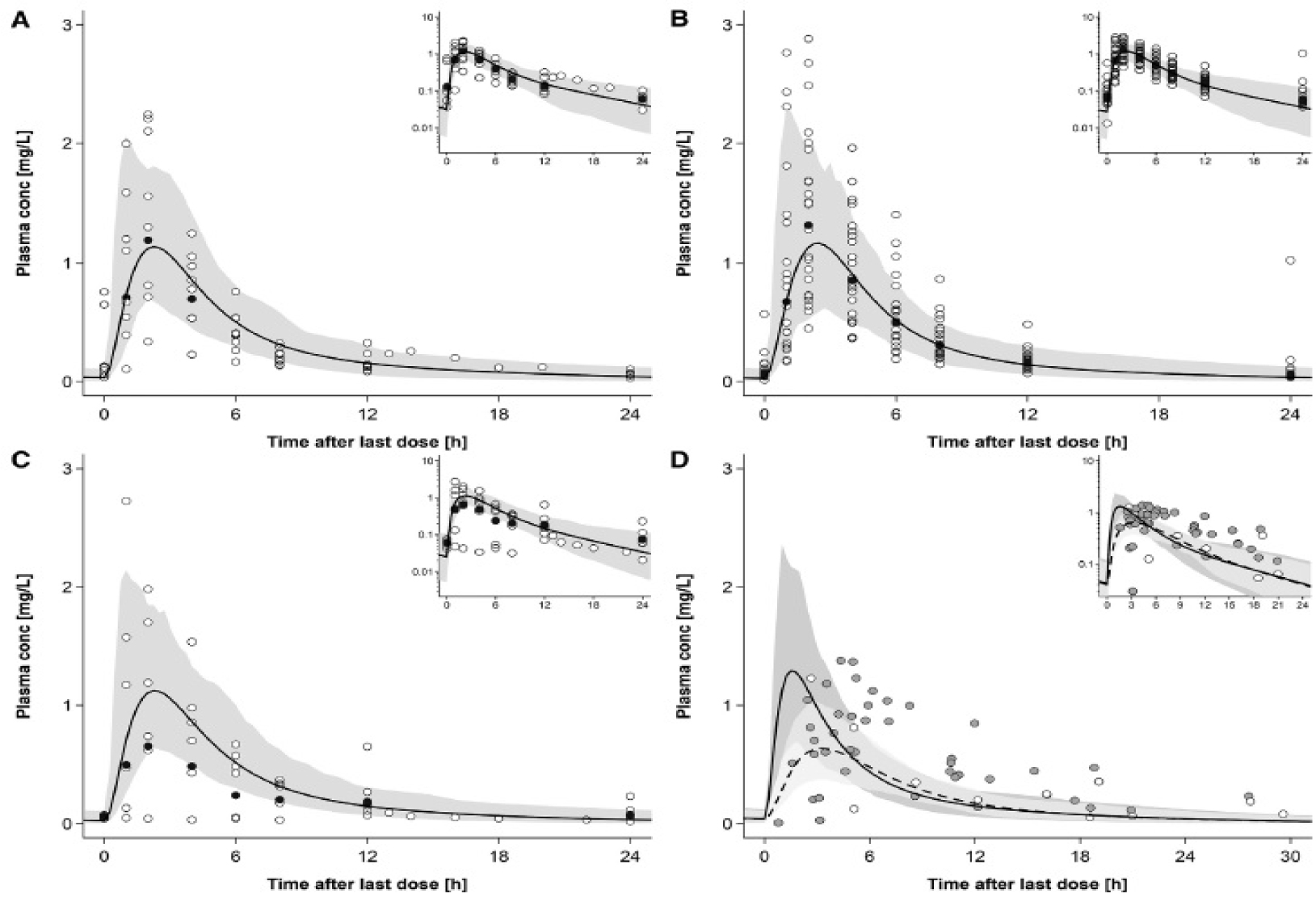
Plasma concentration-time profiles of emtricitabine following oral administration of 200 mg in pregnant women in steady state. Semi-log scale figures are given as inset figures in the top right corners. **A**, Pregnant women with a gestational age of 23-30 weeks. **B**, Pregnant women with a gestational age of 31-35 weeks. **C,** Pregnant women with a gestational age of 36-42 weeks. D, represents individual concentration data in maternal plasma and umbilical vein. ^21,2^

### Non-pregnant and Pregnant PBPK models

#### Dolutegravir and Raltegravir

##### Dolutegravir

The simulated of plasma concentration-time profiles of DTG in non-pregnant populations result from simulation following the administration of a 50 mg tablet once daily in a fed state, mirroring the dosing regimen employed in pregnant women (Fig. 5). Additionally, the figure shown simulated plasma concentration-time profiles for various dosing regimens. Table 1 provides the ratios of simulated to observed pharmacokinetic (PK) parameters in non-pregnant subjects, presenting both the absolute simulated and observed values for these parameters.

**Figure 5:**
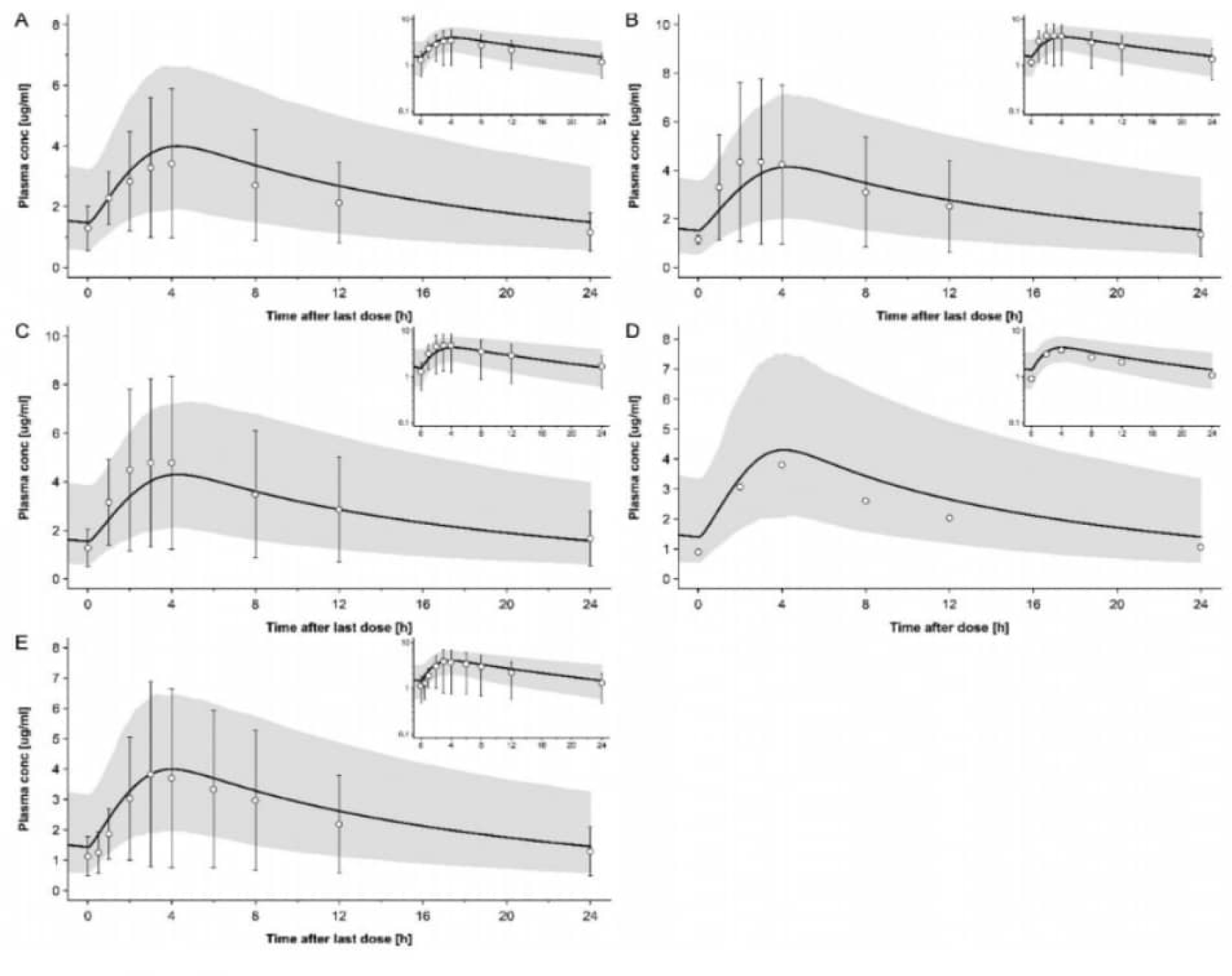
Plasma concentration-time profiles of dolutegravir following oral administration once a day of 50 mg in fed state in non-pregnant subjects.^31,34,35,36^

**Table 1.** provides a comparative analysis between the observed in vivo pharmacokinetic (PK) parameters and those derived from the simulated concentration-time profiles for Emtricitabine, Acyclovir, Dolutegravir, and Raltegravir, respectively.

| Name of Antiviral drug and population condition | AUC<sub>0-t</sub> [mg·h/L]<br>Simulated or Predicted/Observed (Ratio) | C<sub>max</sub> [mg/L]<br>Simulated or Predicted/Observed (Ratio) | t<sub>max</sub> [h]<br>Simulated or Predicted/Observed (Ratio) |
| --- | --- | --- | --- |
| **Emtricitabine** |  |  |  |
| Nonpregnant women |  |  |  |
| Bapuji et al<sup>20</sup> | 10.60/11.20 (0.95) | 3.00/2.01 (1.49) | 0.90/0.78 (1.15) |
| Zong et al, study 1<sup>19</sup> | 9.67/11.20 (0.86) | 2.46/2.13 (1.16) | 0.95/1.01 (0.94) |
| Zong et al, study 2<sup>19</sup> | 9.62/11.11 (0.87) | 2.36/2.17 (1.09) | 0.95/1.25 (0.76) |
| Blum et al<sup>18</sup> (steady state) | 10.70/10.33(1.04) | 1.33/1.64 (0.81) | 3.00/3.07 (0.98) |
| Zong et al, study 3<sup>19</sup> (steady state) | 9.62/9.67 (0.99) | 2.44/2.11 (1.16) | 1.00/1.02 (0.98) |
| Pregnant women |  |  |  |
| GA 23-30 weeks<sup>21</sup> (steady state) | 7.41/7.50 (0.99) | 1.14/1.19 (0.96) | 2.25/2.00 (1.13) |
| GA 31-35 weeks<sup>21</sup> (steady state) | 7.35/6.45 (1.14) | 1.17/1.32 (0.89) | 2.50/2.00 (1.25) |
| GA 36-42 weeks<sup>21</sup> (steady state) | 7.27/4.15 (1.75)<sup>a</sup> | 1.12/0.66 (1.70)<sup>a</sup> | 2.25/2.00 (1.13)<sup>a</sup> |
| **Acyclovir** |  |  |  |
| Nonpregnant women |  |  |  |
| Intravenous injection<sup>12</sup> |  |  |  |
| Study group A (2.5 mg/kg) | 13.7/11.5 (1.19) |  |  |
| Study group B (5.0 mg/kg) | 22.8/23.3 (0.98) |  |  |
| Study group C (10 mg/kg) | 38.8/37.6 (1.03) |  |  |
| Study group D (15 mg/kg) | 56.9/44.9 (1.27) |  |  |
| Oral administration<sup>13</sup> |  |  |  |
| Suspension | 2.98/2.51 (1.19) | 0.640/0.543 (1.18) | 1.60/1.50 (1.07) |
| Tablet | 3.09/2.50 (1.24) | 0.669/0.590 (1.13) | 1.65/1.75 (0.94) |
| Pregnant women |  |  |  |
| GA 36 weeks<sup>12</sup> | 2.00/2.11 (0.95) | 0.574/0.630 (0.91) | 1.65/2.00 (0.83) |
| GA 38 weeks<sup>12,13</sup> (steady state) | 2.84/3.05 (0.93) | 0.726/0.791 (0.92) | 1.65/1.50 (1.10) |
| **Dolutegravir** |  |  |  |
| Nonpregnant women |  |  |  |
| Castellino study<sup>28</sup> | 34.0/35.9 (0.95) | 2.29/2.53 (0.86) | 0.95/0.50 (1.90) |
| Dooley study cohort 1<sup>29</sup> (steady state) | 39.0/36.1 (1.08) | 2.91/2.65 (1.10) | 2.25/1.5 (1.50) |
| Dooley study cohort 2<sup>29</sup> (steady state) | 41.1/42.1 (0.98) | 2.96/2.91 (1.02) | 2.20/2.00 (1.10) |
| Ford study<sup>34</sup> (steady state) | 63.1/52.5(1. 20) | 4.00/3.43 (1.17) | 4.25/4.00 (1.06) |
| Johnson2014, Cohort 1 (steady state)<sup>35</sup> | 65.2/71.9 (0.91) | 4.14/4.35 (0.95) | 4.30/3.00 (1.43) |
| Johnson2014, Cohort2 (steady state)<sup>35</sup> | 67.4/71.9 (0.94) | 4.30/4.78 (0.90) | 4.30/3.50 (1.23) |
| Song2012_high_fat<sup>32</sup> | 66.1/83.6 (0.79) | 2.97/4.19 (0.71) | 4.90/5.00 (0.98) |
| Song2012_low_fat<sup>32</sup> | 59.8/66.7 (0.90) | 2.83/3.81 (0.74) | 4.00/3.00 (1.33) |
| Song2012_moderate_fat<sup>32</sup> | 64.7/71.0 (0.91) | 2.94/3.86 (0.76) | 4.75/4.00 (1.19) |
| Song2016_moderate_fat_meal<sup>31</sup> (steady state) | 62.2/55.4 (1.12) | 3.99/3.83 (1.04) | 4.00/3.00 (1.33) |
| Song2013 study<sup>30</sup> b | 47.2/40.3 (1.17) | 1.82/1.90 (0.96) | 2.50/3.00 (0.83) |
| Weller study<sup>33</sup> | 44.1/37.1 (1.19) | 1.89/1.84 (1.03) | 2.40/2.50 (0.96) |
| Wang2019<sup>36</sup> | 63.8/51.62 (1.23) | 4.30/3.81(1.13) | 4.10/4.00 (1.03) |
| Pregnant women |  |  |  |
| 2nd trimester (steady state) | 34.70/42.38 (0.82) | 2.77/3.00 (0.92) | 4.20/2.00 (2.10) |
| 3rd trimester (steady state) | 31.91/47.59 (0.67) | 2.57/3.00 (0.86) | 4.20/4.00 (1.05) |
| **Raltegravir** | See OSP GitHub | See OSP GitHub | See OSP GitHub |
| Non-pregnant women |  |  |  |
| Markowitz2006<sup>54</sup> | 8.86/7.96 (1.11) | 3.04/2.24 (1.36) | 0.80/1.00 (0.80) |
| Iwamoto2009<sup>57</sup> | 8.66/4.90 (1.77) | 3.10/1.28 (2.42) | 0.75/1.50 (0.50) |
| Rhee2014<sup>55</sup> | 9.13/8.53 (1.07) | 3.11/2.22 (1.40) | 0.75/2.00 (0.38) |
| Wenning2009<sup>56</sup> | 8.99/12.25 (0.73) | 3.12/3.82 (0.81) | 0.75/1.50 (0.50) |
| Brainard2011_fasted b<sup>58</sup> | 9.66/6.47 (1.49) | 3.42/1.59 (2.15) | 0.75/2.00 (0.38) |
| Brainard2011_high fat b<sup>58</sup> | 8.83/ 11.37 (0.78) | 1.48/1.59 (0.93) | 2.45/2.00 (1.23) |
| Brainard2011_moderate_fat b<sup>58</sup> | 8.86/6.44 (1.38) | 1.54/0.74 (2.08) | 2.20/4.00 (0.55) |
| Brainard2011_low_fat b<sup>58</sup> | 9.01/3.39 (2.66) | 1.76/0.59 (2.98) | 1.95/3.50 (0.56) |
| Taburet2015_moderate_fat_1 c<sup>59</sup> | 8.46/8.24 (1.03) | 1.65/2.03 (0.82) | 2.20/1.00 (2.20) |
| Taburet2015_moderate_fat_2 c<sup>59</sup> | 8.95/ 11.00 (0.81) | 1.54/2.77 (0.56) | 2.20/2.00 (1.10) |
| Pregnant women |  |  |  |
| 2nd trimester (steady state) | 4.10/3.90 (1.05) | 0.834/0.67 (1.22) | 2.55/2.00 (1.28) |
| 3rd trimester (steady state) | 3.71/4.44 (0.84) | 0.763/0.85 (0.89) | 2.50/2.00 (1.25) |

Figure 6 illustrates the anticipated plasma concentration-time profiles of DTG during the middle and final stages of pregnancy. In Figure 7, the Goodness-of-Fit (GOF) plot displays model-predicted DTG plasma concentrations in both non-pregnant and pregnant women. Tab. 4 provides absolute values and ratios of predicted to observed AUC 0-24, Cmax, and tmax in the pregnant populations. The model adequately captured variability, with the predicted 5th – 95th percentile range encompassing 76% of all observed concentration values in the mid-pregnancy stage and 69% in the final stage of pregnancy.

**Figure 6:**
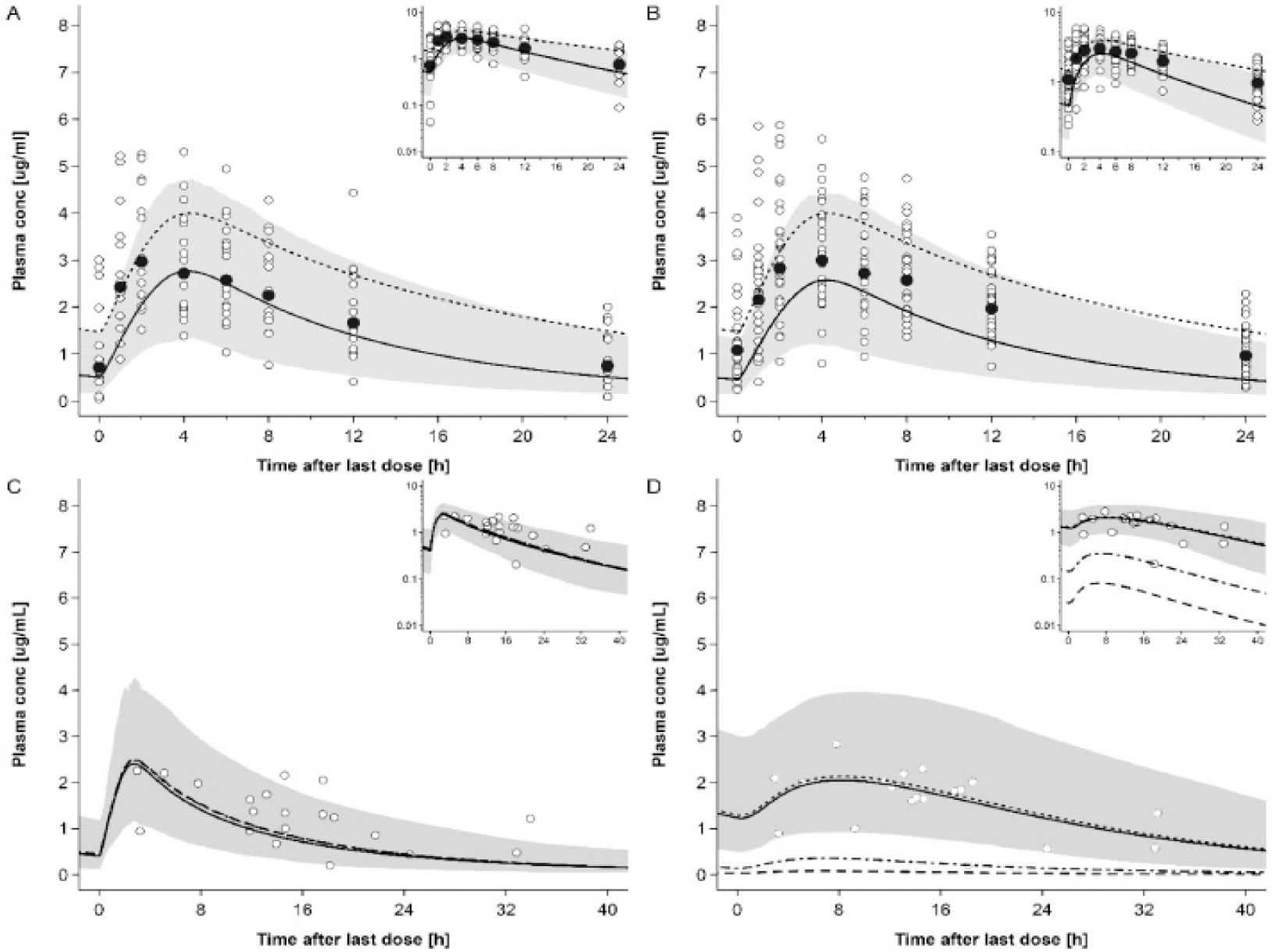
Plasma concentration-time profiles of dolutegravir following oral administration of 50 mg once a day in pregnant women. Semi-log scale figures are given as inset figure in the top right corners.^40^

**Figure 7:**
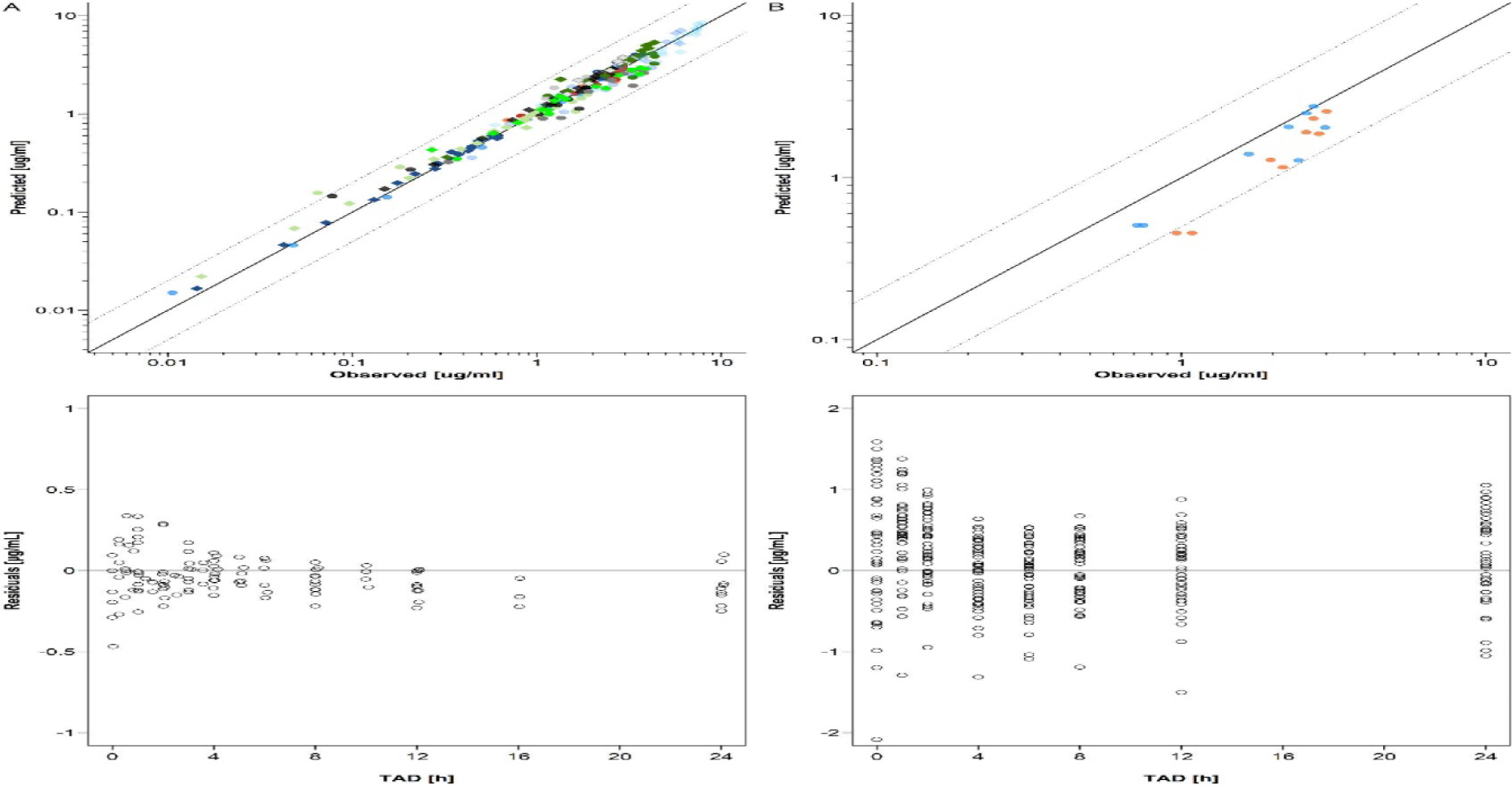
Goodness-of-fit (GOF) and residuals vs time plots of dolutegravir concentrations in non-pregnant subjects (A) and pregnant women (B). **A panel**: Upper plot: GOF plot of geometric mean dolutegravir concentrations in non-pregnant populations. **B panel**: Upper plot: GOF plot of dolutegravir in pregnant population.^28-36^

##### Raltegravir

The simulated plasma concentration-time profiles of RAL in non-pregnant populations following administration of 400 mg tablet BID in fed state (i.e. the same dosing regimen as in pregnant women) are shown in Fig. 8.

**Figure 8:**
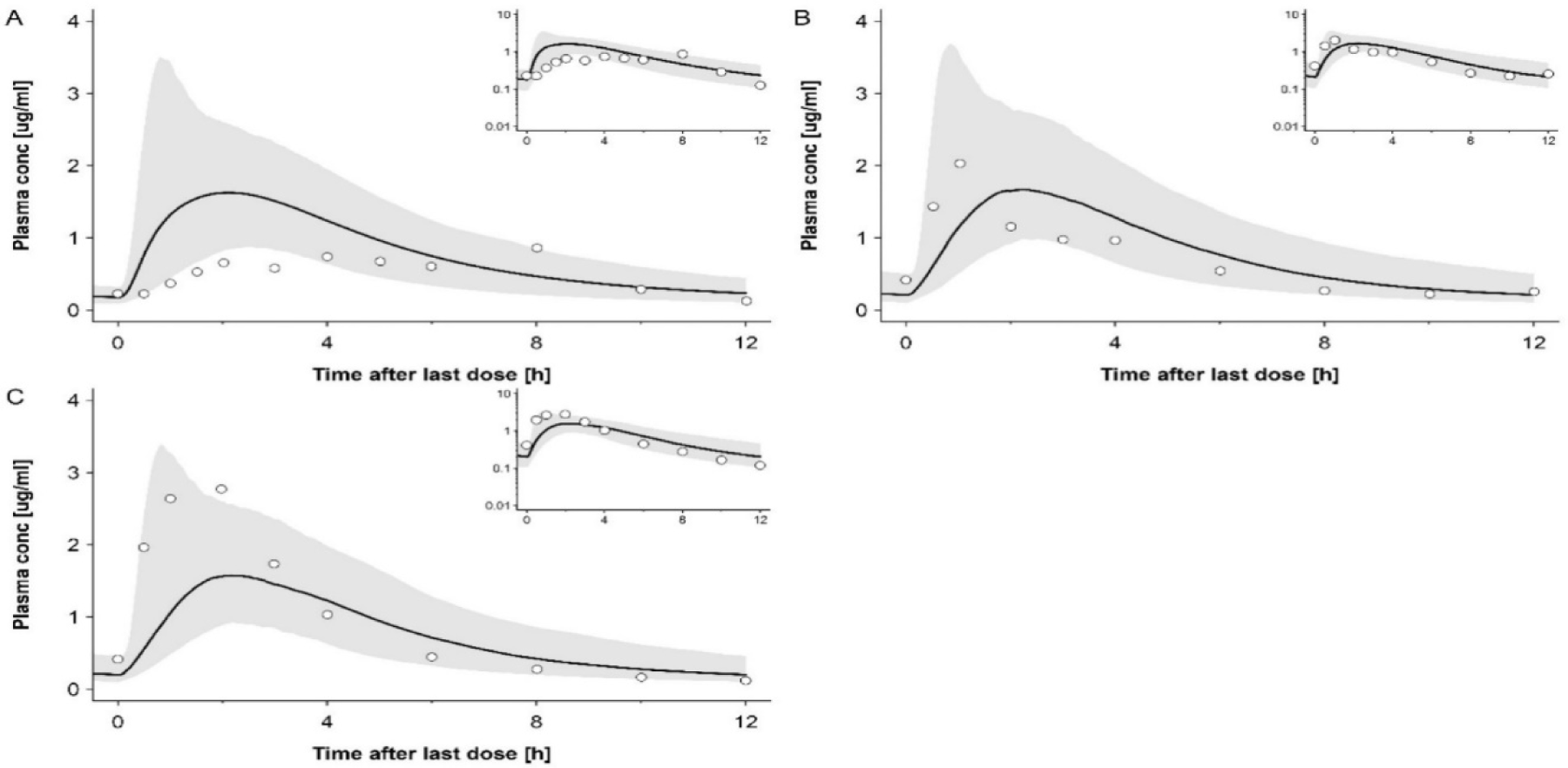
Plasma concentration-time profiles of raltegravir following oral administration twice a day of 400mg with moderate fat meal in non-pregnant subjects.^54,55^

The predicted RAL plasma concentration-time profiles in the 2nd and 3rd trimesters of pregnancy are shown in Fig. 9

**Figure 9:**
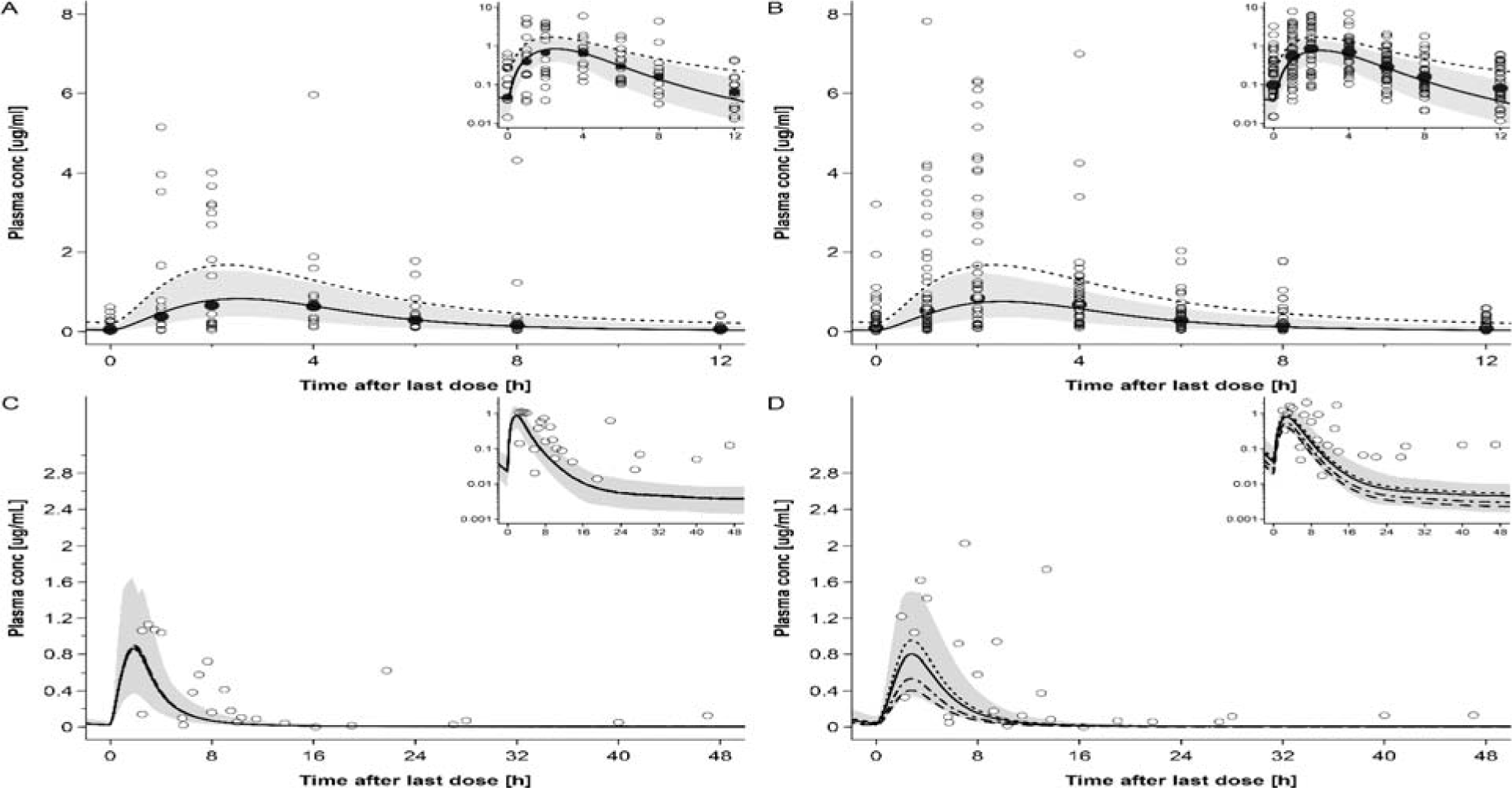
Plasma concentration-time profiles of raltegravir following oral administration of 400 mg twice a day in pregnant women in steady state. Semi-log scale figures are given as inset figure in the top right corners. **A**: raltegravir 400 mg twice a day in pregnant women in 2nd trimester. **B**: raltegravir 400 mg twice a day in pregnant women in 3rd trimester. **C**: raltegravir 400 mg twice a day in pregnant women with an average gestational age of 38 weeks at delivery, represent individual concentration data in the maternal plasma. **D**: raltegravir 400 mg twice a day in pregnant women with an average gestational age of 38 weeks at delivery, represent individual concentration data in the umbilical vein.^41^

Fig. 10 shows the GOF plot for the model-predicted RAL plasma concentrations in non-pregnant and pregnant women with the residuals versus time.

**Figure 10:**
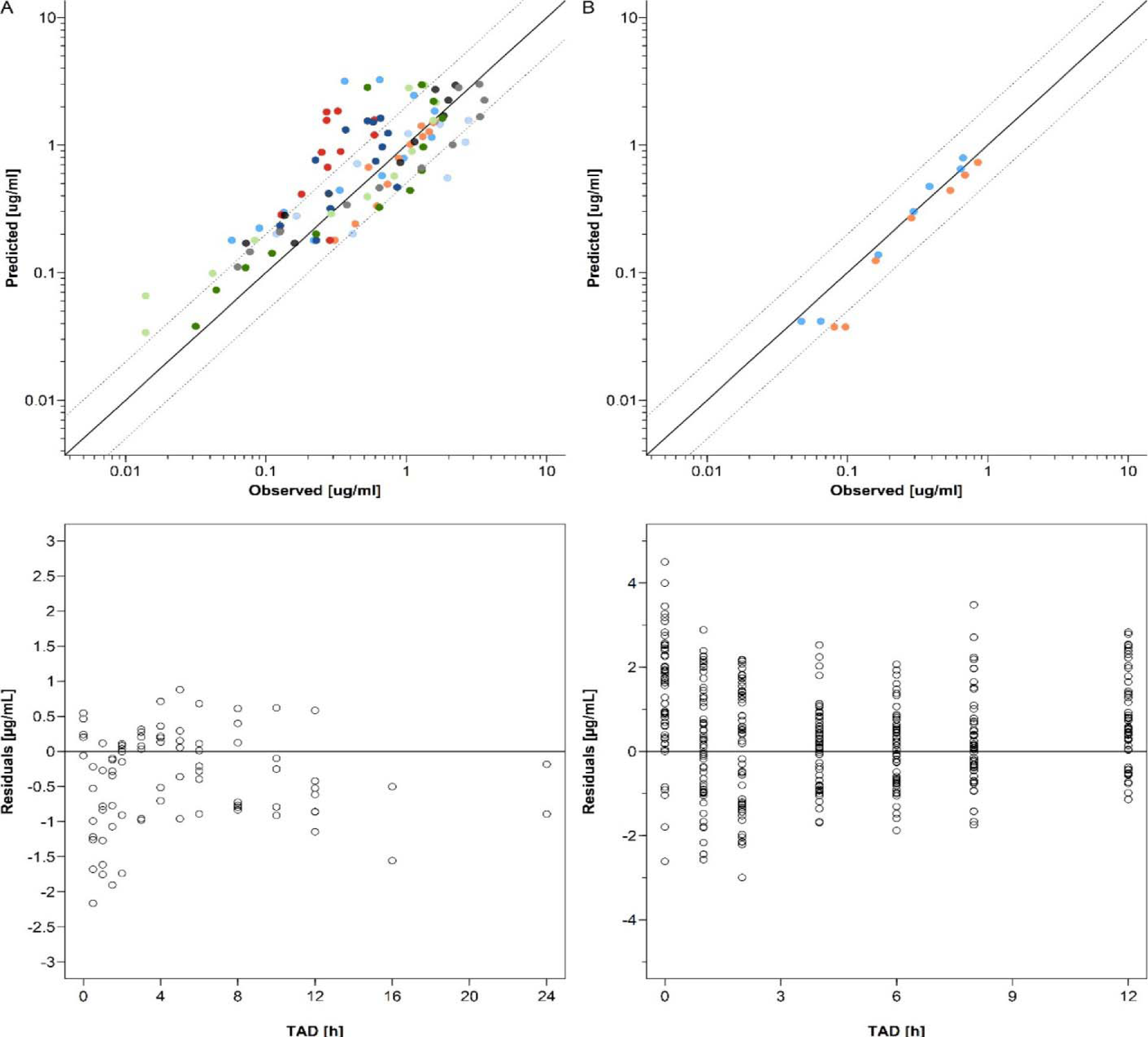
Goodness-of-fit and residuals vs time plots of raltegravir concentrations in non-pregnant subjects (A) and pregnant women (B). **A panel**: Upper plot: GOF plot of geometric mean dolutegravir concentrations in non-pregnant populations. **B panel**: Upper plot: GOF plot of raltegravir in pregnant population.^54, 55^

## Discussion

The present research study focused on the development and evaluation of maternal-fetal PBPK models for four drugs: Emtricitabine, Acyclovir, Dolutegravir, and Raltegravir. In order to assess the accuracy of these models, in vivo data was utilized, including maternal and fetal concentration data collected at different stages of pregnancy. The study found that, overall, the maternal pharmacokinetics (PK) were well-predicted by the developed models. Specifically, the observed values for Cmax, tmax, and AUC_0_-tlast were predicted within a 25% margin of error, as shown in Table 1. However, it should be noted that the description of interindividual variability was less satisfactory, particularly for Acyclovir. This limitation is commonly observed in current PBPK models. In the presented models, changes in drug distribution were primarily driven by increases in the volume of blood plasma and other tissues, such as fat tissue. Additionally, changes in the drug’s fraction unbound also played a role. It is worth mentioning that no PK data following intravenous administration of the modeled drugs were reported, which complicates the evaluation of predicted drug distribution. Nonetheless, previous studies on pregnancy PBPK models for intravenously administered drugs, such as cefazolin, cefradine, cefuroxime, and acetaminophen, have indicated that the disposition kinetics at various stages of pregnancy were adequately predicted. This provides some confidence in the validity of the presented maternal-fetal PBPK models.^24,26^The nonpregnant model presented for intravenous administration was developed using pharmacokinetic (PK) data from men. However, it is important to note that this approach may have resulted in an underestimation of acyclovir clearance when applied to pregnant women. In the study protocol, it was observed that atazanavir was concomitantly administered in 11 patients following the intake of a light meal (∼360 kilocalories). This suggests that, at least in these patients, emtricitabine was taken in a fed state. As a result, the incorporation of a light meal intake was implemented in all maternal-fetal PBPK models. This adjustment led to a decrease in C_max_ (maximum concentration) and an increase in tmax (time to reach maximum concentration) compared to the simulations in nonpregnant women. However, AUC_0-t_ (area under the curve from time zero to the last measurable concentration) was not affected and remained virtually identical between predictions for the fasted and fed states in pregnant populations. These findings indicate that there may be a negative impact of food on emtricitabine PK. However, since the overall exposure is not affected, this food effect is likely of no clinical significance. It is worth noting that a small fraction of acyclovir (8.5% to 14.1%) is metabolized through the action of aldehyde dehydrogenase 2 (ALDH2). The available evidence suggests that a similar conclusion can be drawn for emtricitabine regarding its metabolism, although the specific enzyme involved in this process is not yet known. Emtricitabine undergoes two main metabolic pathways: oxidation of the thiol moiety, leading to the formation of 3r-sulfoxide diastereomers, and conjugation with glucuronic acid to form 2r-O-glucuronide. Notably, these metabolic reactions do not seem to be catalyzed by the cytochrome P450 enzyme system, as mentioned in the presented PBPK models. Interestingly, the PBPK models presented in this study indicate that the metabolic clearance of both acyclovir and emtricitabine remains unchanged during pregnancy. It is worth noting that both emtricitabine and acyclovir are substrates of renal drug transporters, implying that renal excretion plays a significant role in their elimination from the body. Emtricitabine has been found to be a substrate for efflux transporters such as MATE1, MATE2, and MRP1. However, the specific influx transporter for emtricitabine has not yet been identified. On the other hand, acyclovir is a substrate for multiple transporters, including OAT1, OAT2, OAT3, OCT1 for influx, and MATE1 and MATE2 for efflux. Unfortunately, there is currently limited evidence regarding the impact of pregnancy on the expression of these transporters. In the maternal-fetal PBPK models presented, the predicted increase in total renal clearance can be attributed to various factors, including the rise in glomerular filtration rate, kidney volume, renal blood flow, and fraction unbound. Similar to a previous study, a univariate sensitivity analysis was conducted on these parameters. Specifically, the plasma concentration-time profile for acyclovir and emtricitabine was simulated while these factors in the model were either kept constant at the nonpregnant value or adjusted to the pregnant value. The analysis conducted revealed that the observed increase in total renal clearance during pregnancy could not be solely explained by the rise in glomerular filtration rate, fraction unbound, and renal blood flow. Interestingly, it was found that the higher kidney volume played a significant role in predicting the increase in tubular net secretion, which directly influenced the tubular secretion rate and ultimately led to the observed increase in total renal clearance. Currently, there is limited information available on renal physiology and renal transporters in pregnant women, which poses challenges in identifying the specific factors contributing to the observed increase in tubular net secretion clearance. Previous pregnancy PBPK models for emtricitabine have incorporated the observed increases in net secretion clearance of metformin, a well-known substrate of OCT2, or increases in renal plasma flow to inform the rise in tubular net secretion. In the current model, a 52% increase in tubular net secretion was applied in the third trimester. However, further in vitro and in vivo data are required to uncover the underlying physiological mechanism(s) responsible for this clearance increase. In the case of acyclovir, it was observed that the predicted fetal concentrations were relatively insensitive to changes in the transfer constant but were sensitive to changes in the partition coefficient. The PBPK model used in this study slightly underestimated fetal concentrations of acyclovir due to a smaller partition coefficient, as determined from ex vivo cotyledon perfusion data, resulting in higher concentrations on the maternal side compared to the fetal side of the placenta. However, it is questionable whether the in vivo partition coefficient for acyclovir is indeed smaller than 1. It is important to note that this study focused on investigating pregnancy-induced alterations in pharmacokinetic (PK) target parameters, and further considerations on appropriate dosing regimens would require incorporating drug pharmacodynamics, which was beyond the scope of this study. It should be acknowledged that the patients included in this study, who were living with HIV, were taking multiple antiviral drugs. The clinical study used for model evaluation during pregnancy involved co-administration of emtricitabine with several other antiretroviral medications. However, it is expected that none of these co-medications interact with emtricitabine. Although no drug-drug interactions were anticipated in this study, the therapeutic effect, such as viral load suppression, represents a combined effect of multiple antiviral drugs, making the development of a mechanistic pharmacodynamic model complex. Regarding acyclovir, the median minimum concentration (C_min_) and area under the curve (AUC) predicted throughout pregnancy consistently remained above the reported inhibitory concentration (IC50). Similarly, for emtricitabine, the median Cmin and AUC predicted throughout pregnancy were also above the reported target thresholds. However, in the lower percentiles, both Cmin and AUC during late second trimester and early third trimester were predicted to be lower than the IC50 and desired AUC, respectively. The PBPK models developed in this study were based on the population described and have not been adjusted for pharmacogenetic differences or ethnicity. However, it is not expected that there are any significant changes in this regard for emtricitabine and acyclovir. The predicted PK of DTG in pregnant, non-laboring women aligned well with clinical data obtained at different stages of pregnancy. While most PK parameters were accurately predicted, the time to reach maximum concentration (tmax) in the second trimester was slightly overestimated, and the AUCtau was underestimated in the third trimester. Further clinical studies controlling for food intake could help elucidate the reasons behind these observations. The underestimation of AUCtau in the third trimester for DTG was primarily attributed to an overestimation of total body clearance. This, combined with the results from the sensitivity analysis on UGT1A1 induction, suggests that UGT1A1 induction may be lower than initially expected, and changes in the fraction unbound are the main driver of increased total clearance, which is consistent with recent findings in the literature. The PK of RAL in pregnant, non-laboring women was generally well predicted, although variability, particularly in the first hours after drug administration, was underestimated, indicating that the model did not fully capture variability related to drug absorption. UGT1A1 plays a crucial role in the metabolism of both DTG and RAL. While in vitro experiments support an increase in UGT1A1 expression mediated by rising progesterone levels during pregnancy, there is limited information on quantitative changes in UGT1A1 expression in vivo. In this study, UGT1A1 was assumed to be induced by factors of 1.75 in the second trimester and 1.92 in the third trimester based on a previous PBPK model for acetaminophen. Maternal PK of DTG was moderately sensitive to alterations in UGT1A1 expression, while maternal PK of RAL was weakly affected. It is important to acknowledge that the limited clinical data available hindered a comprehensive evaluation of these predictions, and more data, ideally from different tissues such as maternal plasma, placenta, and umbilical vein, are needed to better assess the predictive performance of these models. However, within the limitations of this study, models that incorporated the Poulin & Theil method or the QSAR approach appeared to better predict umbilical vein concentrations compared to other models. This highlights the significance of maternal PK in fetal drug exposure and suggests that the primary elimination pathway in the fetus is transfer across the placenta back to the mother. The concentrations of RAL in maternal plasma and the umbilical vein were particularly underestimated after 12 hours, which corresponds to the dosing interval. One of the reasons for the underestimation of maternal plasma concentrations of RAL at delivery seems to be the fast absorption assumed in the model. While the PBPK models for non-laboring women considered a fed state to reflect the lack of food restrictions in the clinical study, the PBPK model for laboring women assumed a fasted state (as it is unlikely for women to consume food right before labor and delivery). However, it was noted that the PBPK model at delivery still accounted for delayed gastric passage of RAL (and DTG) due to evidence suggesting that gastric emptying and drug absorption from the gastrointestinal tract are slowed during labor. On May 18th, 2018, the US FDA issued a warning letter regarding DTG, stating that it may cause serious birth defects involving the brain, spine, and spinal cord. These preliminary findings were observed in a study conducted in Botswana, where women who received DTG at the time of conception were affected. The exact mechanism behind the teratogenicity of DTG is not yet fully understood, but one hypothesis suggests that it interferes with folic acid binding to the folate receptor α, leading to reduced levels of folic acid in the fetus. Since folic acid is crucial for neural tube development, a decrease in folic acid levels could potentially result in neural tube defects in the fetus. In vitro results presented by Zamek et al. indicate that free DTG concentrations of approximately 37 µM correspond to a 36% inhibition of the folate receptor α. To provide an in vivo context, the PBPK model was extrapolated to the 6th gestational week, assuming a 33% induction of UGT1A1 in the first trimester. Unbound DTG concentrations were then predicted in the maternal blood of the placenta, with a maximum predicted concentration of 0.06 µM at steady state during the 6th gestational week. Using an Emax model fitted to the data reported by Zamek et al., where Emax was 1.0 and EC50 was 1276 µM, this DTG concentration corresponds to an inhibition of the folate receptor α by approximately 7%. However, it is important to note that these predicted concentrations cannot be evaluated due to the lack of clinical data, and therefore should not be used to guide dosing decisions. This example highlights the potential theoretical contribution of PBPK modeling in supporting decision-making regarding the use of DTG during pregnancy. However, it also emphasizes the need for clinical data to validate and enhance the reliability of model-based predictions.

## Conclusions

In summary, the developed maternal-fetal PBPK models have successfully predicted the pharmacokinetic profiles of Emtricitabine, Acyclovir, Dolutegravir, and Raltegravir at various stages of pregnancy. This enhances confidence in utilizing one of the key strengths of PBPK analyses, which is the ability to extrapolate drug pharmacokinetics from well-characterized populations of healthy adults to the unique population of pregnant women. The investigation into pregnancy-induced changes in pharmacokinetic target parameters confirms the appropriateness of current dosing regimens for acyclovir and emtricitabine, at least for the average pregnant patient. The presented model strengthens the confidence in such models, which is crucial when applying them to inform the design of clinical trials for drugs with similar pharmacokinetic characteristics in pregnant women. Given the limited participation of pregnant women in clinical trials, PBPK modeling can serve as a valuable tool to supplement the understanding of pharmacokinetics in cases where clinical data is sparse or unavailable. While these models should not be seen as a substitute for clinical trials, they contribute to a broader and mechanistic understanding of pharmacokinetics, with the potential to enhance drug safety and efficacy for both the mother and the fetus.

## Data Availability

Data Availability from Open Systems Pharmacology provides research data availability through its platforms like PK-Sim and MoBi. These tools enable researchers to build, simulate, and analyze quantitative systems pharmacology models. The availability of these resources promotes data sharing and collaboration, enhancing the reproducibility and transparency of research. By offering access to comprehensive pharmacokinetic and pharmacodynamic modeling data, Open Systems Pharmacology supports the development of new drugs and the advancement of pharmacological research.

https://github.com/Open-Systems-Pharmacology

